# Thiamine alone rather than in combination with ascorbic acid is associated with improved survival in septic shock

**DOI:** 10.1101/2023.04.14.23288576

**Authors:** David Legouis, Aimad Ourahmoune, Sebastian Sgardello, Frederic Sangla, Gilles Criton

## Abstract

**Background:** Sepsis and septic shock are common causes of ICU admission with devastating outcomes. Adjunctive therapies are urgently needed, and the use of high dose of vitamin B1 and C have recently gained interest. However, on the basis of a perceived possible synergic effect, most trials have never tested the combination of thiamine and ascorbic acid, with a separate assessment of the effect of each individual component. In this context, while the association of thiamine and ascorbic acid was not found to improve survival rates, potentially harmful effects were found when administering ascorbic acid alone. We have conducted a retrospective cohort study, comparing ICU mortality of septic shock patients receiving standard treatment, thiamine alone or a combination of thiamine and ascorbic acid.

**Results:** A total of 1800 patients were included, 1260 receiving standard care, 436 receiving only thiamine and 104 patients receiving a thiamine / ascorbic acid combination. Using doubly robust estimation of the treatment effect, combining propensity score weighting and variables adjustment, we found thiamine alone to be associated with a decrease in ICU mortality compared to the use of a thiamine / ascorbic acid combination (Hazar Ratio equal to 0.60, 95% Confidence Interval [0.36;0.99], p=0.048).

**Conclusions:** In septic shock patients, administration of thiamine is associated with improved ICU mortality when used alone rather than when associated with ascorbic acid. This result strengthens the evidence showing a lack of effectiveness of the ascorbic acid / thiamine combination reported in recent randomized controlled trials. Furthermore, it argues in favor of the need for further trials investigating the effect of thiamine in septic ICU patients as an adjunctive therapy.

## Background

Sepsis is a common and life-threatening condition related to a deregulated host response to infection, leading to organ dysfunction[1–4]. Depending on which studies are taken into account, its related ICU mortality can reach 50% [1,2,5–8]. In addition, the severity of illness of septic ICU patients seems to be increasing over the last decade, with a higher proportion of patients in septic shock and an increase in the need for renal replacement therapy[9].

To this day, sepsis and septic shock management relies on antimicrobial administration, fluid resuscitation and control of the source of the infection and use of vasopressors when appropriate[10– 12]. In 2017, an observational before/after study reported a dramatic improvement in septic patients’ outcomes when using a combination of hydrocortisone, ascorbic acid and thiamine (HAT therapy) [13]. The HAT therapy or metabolic cocktail resuscitation was further assessed in several randomized controlled trials (RCTs) with none reaching a statistical significance for their respective primary outcomes. A meta-analysis of these studies conducted by our group found a modest improvement in duration of organ dysfunction but no survival benefit with the use of HAT therapy[14].

One of the rationales supporting the use of HAT therapy was the perceived positive synergic effect between its constituents[13], which has, however, never been proven. Thus, unexpected interactions between thiamine and ascorbic acid may otherwise or cancel out convolute effects, limiting the ability to assess the potential benefit of each compound separately. In this context, and despite existing trials on HAT therapy, questions about the effects of thiamine and ascorbic acid administered in monotherapy remain.

In this study, we hypothesize that thiamine may exert different individual effects in ICU patients with septic shock when it is combined or not with ascorbic acid.

## Material and Methods

### Aims of the study

This study aims at comparing the ICU mortality of patients with septic shock receiving standard treatment, standard treatment with thiamine supplementation or standard treatment with the administration of a combination of thiamine and ascorbic acid.

### Study design

We conducted a single-center retrospective cohort study with multinomial propensity score weighting analyses. This study was carried out in accordance with the principles outlined in the Declaration of Helsinki and was approved by the local ethical committee for human studies of Geneva, Switzerland (Commission Cantonal d’Ethique de la Recherche, CCER 2023-00147).

### Population Study

All adult patients (>18 years of age) admitted to the ICU of the Geneva University Hospitals between January 2012 and August 2022 were screened. Among them, patients with a final diagnosis of septic shock, were included.

### Groups of Patients

Three groups of patients were defined, according to the treatment they received within the first 48 hours following ICU admission. The control group involved patients receiving only standard treatment for septic shock. Thiamine (B1) and thiamine/ascorbic acid groups (AT) included patients who received standard treatment for septic shock with the addition of thiamine (B1), and patients who received a combination of thiamine and ascorbic acid (AT).

### Treatment Administration

The standard treatment of septic shock was conducted according to international guidelines[12]. Use of thiamine and ascorbic acid was not protocolized in our unit but was left to the discretion of the attending physician. The combination of thiamine and ascorbic acid was used as of 2017 following Marik’s publication [13], while the use of thiamine alone progressively increased over time (**Supplemental Figure 1**). In this context, the date of ICU admission was included as a potential confounding factor in the propensity score.

### Statistics

Baseline characteristics were expressed as median (25-75^th^ percentiles) or absolute and relative (%) frequency if categorical. They were compared using Mann Whitney or a Fisher’s Exact test depending on their class. For downstream analyses (*i*.*e*., propensity score weighting analyses), missing data were imputed using the missForest R package[15], which uses a random forest trained on the observed values to predict the missing values. Previous research established evidence in favor of applying imputation for missing data to improve the accuracy of modelling[16–18]. Among the available methods, multiple imputation using random forest provides several benefits. It handles both numerical and categorical variables without the need for prior preprocessing as no assumption of features relationships are made. It is also robust to multicollinearity and noisy data since it includes built in features selection. Finally, it can fit a nonlinear relationship. Altogether, it emerged as an optimal strategy, that was shown to outperform other methods, although it is limited by the very high computational overhead and the large amounts of memory required[15,18,19].

For the estimation of the multinomial propensity score, we took advantage of the twang package[20]. This package was originally developed to perform propensity score station and weighting using generalized boosted regression. These models are flexible machine learning approaches which are able to deal with non-linearities and interactions across the included variables. The algorithm implemented in twang selects the optimal complexity of the model that achieves the best balance among treatment groups. This package was further extended to handle more than two treatment conditions through the multinomial propensity score[21]. We used this package with default parameters. The estimated estimand was the average treatment effect on the treated (ATT), and the reference treated group was the one receiving the combination of thiamine and ascorbic acid. The following variables were considered as potential confounders:

- year of ICU admission
- administration of hydrocortisone.
- Demographic data and comorbidities (age, sex, chronic pulmonary disease, hypertension, congestive heart failure, diabetes mellitus, renal and liver diseases).
- Severity of illness at ICU admission (saps score, bilirubin and arterial base excess levels, white blood cells and platelets counts, estimated GFR and median heart rate in the first 24 hours).
- Need for organ support therapy within 24 hours of ICU admission (cumulative dose of infused norepinephrine, need for renal replacement therapy, invasive mechanical ventilation, and extracorporeal membrane oxygenation).

We first checked the model run for a sufficiently large number of iterations to minimize the stopping rule while avoiding overfitting. As twang generated 4 sets of weights, each one corresponding to a specific stopping rule, we selected the one which achieved the greatest mean reduction of both the Kolmogorov-Smirnov statistics and the absolute standardized mean difference. The balance of the confounding factors between the three groups was assessed by the absolute standardized mean difference before and after weighting, with a value lower than 25% being considered as sufficient to support the assumption of balance between groups[22].

The optimal propensity score weights were extracted using the get.weights command and were further used in a weighted survival cox model to estimate the treatment effect. We ensured that our model met the proportional hazard assumptions[23]. Additionally, variables displaying absolute standardized mean difference above 0.25 after weighting were added in the model to correct for imbalance, a procedure called doubly robust estimators [24–26].

All analyses were performed using R software. P-values were two-tailed and a value lower than 0.05 was considered significant.

## Results

### Population

Between January 2012 and August 2022, 23131 patients were admitted to the ICU of the Geneva University Hospitals. Among them, 487 patients were not included because they received thiamine or ascorbic acid after more than 48 hours following ICU admission. Of the 22644 remaining patients, 1800 patients admitted for septic shock were considered for the analyses. In this cohort, 1260 patients were included in the control group, 426 in the B1 group and 104 in the AT group. Baseline characteristics of these patients are shown in **Table 1**.

**Table 1.** *Baseline characteristics of patients among groups*. BMI, Body Mass Index; SAPSII, Simplified Acute Physiological Score; APACHE II, Acute Physiology and Chronic Health Evaluation; eGFR, estimated glomerular filtration rate; ECMO, Extra Corporeal Membrane Oxygenation

|  | Control group<br>(N=1260) | B1 group<br>(N=436) | AT group (N=104) | Total<br>(N=1800) | p value |
| --- | --- | --- | --- | --- | --- |
| **Patients' characteristics** |  |  |  |  |  |
| Age (years), median (IQR) | 70 (58, 78) | 65 (57, 74) | 68 (58, 76) | 68 (58, 77) | < 0.001 |
| Sex, male, n (%) | 754 (59.8) | 323 (74.1) | 63 (60.6) | 1140 (63.3) | < 0.001 |
| BMI (kg/m<sup>2</sup>), median (IQR) | 25.4 (22.7, 28.8) | 24.8 (21.7, 28.5) | 25.2 (22.5, 29.4) | 25.2 (22.5, 29.0) | 0.189 |
| SAPS II score, median (IQR) | 51 (39, 65) | 58 (44, 72) | 72 (58, 85) | 53 (41, 69) | < 0.001 |
| APACHE II score, median (IQR) | 25 (19, 31) | 28 (22, 35) | 36 (29, 41) | 26 (20, 33) | < 0.001 |
| Charlson score, median (IQR) | 2 (1, 4) | 2 (1, 4) | 2 (1, 5) | 2 (1, 4) | 0.546 |
| **Underlying diseases** |  |  |  |  |  |
| Diabetes mellitus, n (%) | 188 (14.9) | 65 (14.9) | 12 (11.5) | 265 (14.7) | 0.691 |
| Hypertension, n (%) | 377 (29.9) | 129 (29.6) | 40 (38.5) | 546 (30.3) | 0.184 |
| Chronic heart failure, n (%) | 192 (15.2) | 61 (14.0) | 10 (9.6) | 263 (14.6) | 0.281 |
| Heart valve Diseases, n (%) | 58 (4.6) | 17 (3.9) | 0 | 75 (4.2) | 0.041 |
| Chronic respiratory disease, n (%) | 73 (5.8) | 31 (7.1) | 9 (8.7) | 113 (6.3) | 0.315 |
| Chronic liver disease, n (%) | 46 (3.7) | 34 (7.8) | 11 (10.6) | 91 (5.1) | < 0.001 |
| Active cancer, n (%) | 161 (12.8) | 52 (11.9) | 16 (15.4) | 229 (12.7) | 0.603 |
| **Cause of ICU admission** |  |  |  |  |  |
| Medical, n (%) | 1167 (92.6) | 412 (94.5) | 97 (93.3) | 1676 (93.1) | 0.971 |
| Surgical, n (%) | 93 (7.4) | 24 (5.5) | 7 (6.7) | 124 (6.9) | 0.466 |
| **At ICU admission** |  |  |  |  |  |
| pH, median (IQR) | 7.36 (7.29, 7.41) | 7.35 (7.27, 7.40) | 7.29 (7.24, 7.37) | 7.35 (7.28, 7.41) | < 0.001 |
| Base excess, median (IQR) | -5.2 (-8.5, -1.90) | -5.9 (-9.4, -2.6) | -7.7 (-10.7, -4.3) | -5.6 (-8.8, -2.2) | < 0.001 |
| Lactate (mmol/l), median (IQR) | 1.6 (1.0, 3.0) | 2.0 (1.1, 4.0) | 2.8 (1.8, 4.6) | 1.8 (1.1, 3.3) | < 0.001 |
| Hemoglobin (mg/l), median (IQR) | 101 (87, 118) | 104 (88, 124) | 104 (86, 122) | 102 (87, 119) | 0.058 |
| Platelets (G/l), median (IQR) | 171 (107, 252) | 149 (89, 224) | 155 (84, 209) | 163 (99, 240) | < 0.001 |
| White blood cells (G/l), median (IQR) | 12.1 (6.7, 18.3) | 11.8 (6.5, 18.4) | 12.2 (3.8, 20.9) | 12 (6.5, 18.4) | 0.829 |
| Bilirubin (μmol/l), median (IQR) | 15 (9, 26) | 18 (10, 32) | 15.5 (9.8, 34) | 15 (9, 27) | 0.006 |
| eGFR (mL/min/1.73 m<sup>2</sup>), median (IQR) | 44 (25, 78) | 47 (26, 81) | 31 (19, 52) | 44 (25, 78) | < 0.001 |
| **During first 24 hours upon admission** |  |  |  |  |  |
| Hydrocortisone treatment, n (%) | 583 (46.3) | 292 (67) | 99 (95.2) | 974 (54.1) | < 0.001 |
| Invasive mechanical ventilation, n (%) | 342 (27.1) | 198 (45.4) | 70 (67.3) | 610 (33.9) | < 0.001 |
| Renal replacement therapy, n (%) | 70 (5.6) | 44 (10.1) | 17 (16.3) | 131 (7.3) | < 0.001 |
| ECMO, n (%) | 3 (0.2) | 2 (0.5) | 2 (1.9) | 7 (0.4) | 0.058 |

Briefly, the three groups (*i*.*e*., control, B1 and AT groups) displayed increasing severity of illness. This includes more severe lactic acidosis, higher SAPS or APACHE score at ICU admission, and a higher incidence of mechanical ventilation and renal replacement therapy within the 24 first hours of ICU admission.

The median daily delivered doses of thiamine were 304mg (IQR 193, 600) and 400mg (IQR 233, 600) in the B1 and AT groups respectively. The median daily doses of administered ascorbic acid was 4500mg (IQR 1893, 6100) in the AT group. The median duration of thiamine treatment were 2.23 days (IQR1, 5.5) and 2.05 days (IQR 1, 4.8) in the B1 and AT groups respectively. The median duration of ascorbic acid treatment was 1.11 day (IQR1, 2.28) in the AT group.

### Variables Balance

The absolute standardized mean difference before and after weighting, for each confounding factor and each stopping rule is shown in **Figure 1**. The stopping rule searching for minimization of the mean of absolute standardized mean differences (es.mean) displayed the best overall performance and was further used in the weighted analyses. Before weighting, 11 and 9 variables were unbalanced in the control and thiamine alone groups respectively, while only 4 (use of hydrocortisone, 24h-cumulative doe of norepinephrine, mechanical ventilation, and saps) and 1 variable (use of hydrocortisone) still had an absolute standardized mean difference above 0.25 after weighting. The final weighted model was thus adjusted for these confounding factors.

**Figure 1.**
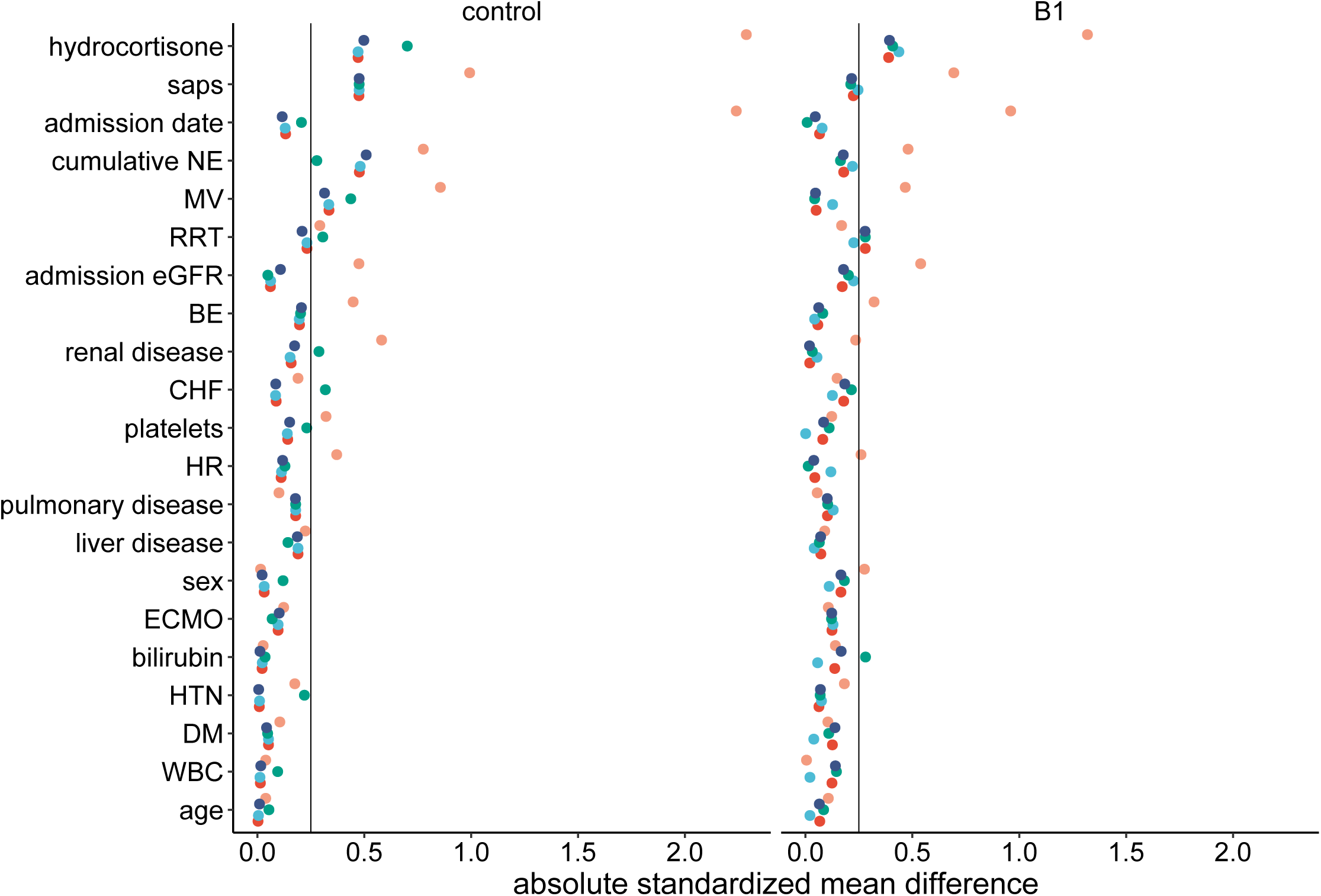
variables balance across groups: Dotplot showing the absolute standardized mean difference each variable included in the propensity score, before and after weighting, for the 4 stopping rules used. NE Norepinephrine; MV Mechanical Ventilation; RRT Renal Replacement Therapy ; eGFR estimated Glomerular Filtration Rate ; BE Base Excess ; CHF Congestive Heart Failure; HR Heart Rate; HTN Hypertension; DM Diabetes Mellitus; WBC White Blood Cell count.

### Treatment Effect

In the crude analysis, control and B1 group displayed a lower ICU mortality compared to the AT group (Hazard Ratio, (HR) equal to 0.68 95% Confidence Interval (CI) [0.47;0.97], p=0.032 and 0.65 [0.44;0.96], p=0.029 respectively). After propensity score weighting (HR=0.60 [0.36;0.99], p=0.047), as well as the use of the doubly robust estimator (HR=0.60 [0.36;0.99], p=0.048), patients from the B1 group showed a decrease in ICU mortality compared to the AT group. The survival curve corresponding to the doubly robust estimation is shown in **Figure 2**.

**Figure 2.**
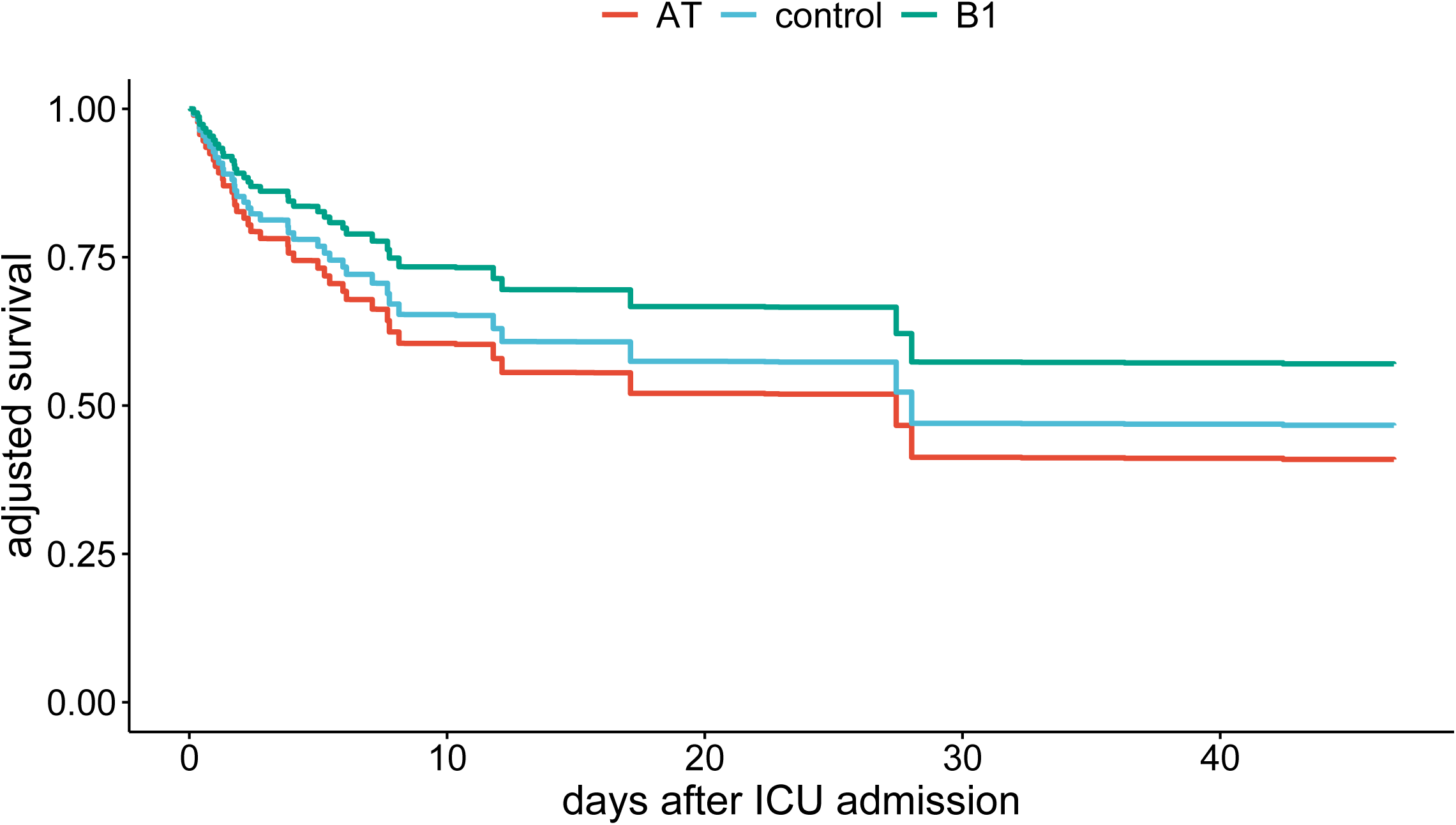
ICU mortality across groups: Survival curves showing the cumulative ICU survival along time across the three groups.

## Discussion

The main finding of our study is an incremental decrease in ICU mortality in AT, control and B1 groups respectively, although only the B1 and the AT groups significantly differ. Interest for vitamin C and B1 as adjunctive therapy for septic shock took a major turn after the publication by Marik et al[13] of an observational before/after study, which reported major improvements of outcomes in septic patients with hydrocortisone, ascorbic acid and thiamine used in combination.

Unlike in the study of Marik et al[13], we did not observe any improvement in ICU mortality in the AT group compared to the control group. This is in line with several recently conducted RCTs which were unable to reproduce the remarkable survival benefit described by Marik et al. [27–33].

Likewise, a recent meta-analysis of these trials confirmed a lack of effect of HAT therapy on mortality despite a slight decrease in 72h SOFA scores[14]. Studies published since have not found any gain in mortality either[34,35].

One of the key concepts of the AT therapy relied on the perceived positive synergic effect between its components[13] although it has never been proven. The negative impact of the addition of ascorbic acid to thiamine observed in our study challenges this assumption. It even raises concerns about potential toxicity of this combination, as patients from AT groups displayed higher ICU mortality compared to both B1 and control groups, although the latter was no longer significant after propensity score weighting. While some authors observed decreased ICU length of stay[36] and mortality[37], improved SOFA score[38] and lowered vasopressor requirements[36,37,39] in septic patients treated with vitamin C, these studies were limited by their small size and their unblinded design. The recent and well conducted CITRIS-ALI trial including 167 patients with sepsis and ARDS reported a lower 28-day mortality, but it was a secondary outcome which was no longer significant when adjusted for multiple comparisons[40]. Neither did two other RCTs find a benefit of ascorbic acid infusion, even in patients with vitamin C deficit at baseline[41,42]. Finally, the potential negative effect associated with the use of ascorbic acid has recently been suggested by the LOVIT trial which is the largest and the most recent multicentric trial. This study involving 35 medical-surgical ICU, enrolled 872 patients in septic shock to receive ascorbic acid (50 mg/kg/6h up to 96 hours) or a placebo. The primary outcome, a composite of death and persistent organ dysfunction at day 28, was significantly higher in the group treated with vitamin C (44.5 versus 38.5%, p=0.01)[43].

The decrease in ICU mortality which we observed in the B1 group compared to the AT patients is in line with a recent RCT. Nandhini et al. randomized 84 septic shock patients in three groups, placebo, thiamine alone (2 mg/kg/8h) and ascorbic acid (50 mg/kg/6h) and found a decrease in ICU mortality in the group treated by thiamine alone (28%) compared to those treated by vitamin C alone (48%) or placebo (60%)[44]. The potential benefit of thiamine alone in comparison with standard treatment in septic patients has already been suggested. A study conducted by Donnino et al., enrolling 88 ICU septic shock patients, compared intravenous infusion of thiamine (200mg/12h) to a placebo. In the predefined subgroup of patients with thiamine deficiency, mortality was decreased in the intervention group (13 versus 46%, p=0.047 for survival analyses)[45]. A similar study was published by Petsakul et al., involving 50 ICU patients in septic shock. It reported a greater reduction of the vasopressor dependency index in patients treated with thiamine[46].

From a biological perspective, the rationale supporting thiamine supplementation exists. Thiamine is a vitamin, acting as a cofactor for the Pyruvate Dehydrogenase Complex (PDH) and the alpha-ketoglutarate dehydrogenase. Both enzymes are involved in the tricarboxylic acid cycle, making thiamine necessary for mitochondrial function and ATP generation[47]. Our group also reported its role in renal gluconeogenesis, a process whose decline is associated with mortality in critical ill patients[48]. In addition, thiamine deficiency is observed in 20-70% of ICU patients[45] and is associated with mortality[49]. Altogether, this argues in favor of further clinical trials evaluating the clinical effect of thiamine alone in ICU patients treated for septic shock.

Our study has several limitations. The first is due to its single-centered design that limits the extent of our findings. The second is that being a retrospective study, results may have been biased due to the presence of confounding factors. However, we used a doubly robust estimation of the treatment effect, combining a propensity score weighting with variables adjustment to achieve balance among groups although we can’t completely rule out the existence of unobserved confounders. The third is related to the use of thiamine and ascorbic acid that was not protocolized in our ICU and may therefore have changed over time. To address this, we included the date of admission in the propensity score and the assessment of this variable was well balanced after weighting.

## Conclusion

We found that the use of thiamine alone within 48 hours after ICU admission in septic shock patients was associated with a lower ICU mortality as compared to its use in combination with ascorbic acid. This finding argues in favor of further, large clinical trials evaluating the effect of thiamine supplementation in septic shock patients as well as highlighting the potentially harmful effects of the thiamine / ascorbic acid combination used in these patients.

## Data Availability

The datasets used and or analyzed during the current study are available from the corresponding author on reasonable request.

## List of Abbreviations

ATT: Average Treatment effect on the Treated
HAT therapy: combination of hydrocortisone, ascorbic acid and thiamine HR: Hazard Ratio
ICU: Intensive Care Unit
RCT(s): Randomized Controlled Trial(s)

## Declarations

### Ethics approval and consent to participate

The study was approved by the local ethical committee for human studies of Geneva, Switzerland (CCER 2023-00147, Commission Cantonale d’Ethique de la Recherche) who waived the need for patient consent. The study was performed according to the Declaration of Helsinki principles

### Consent for publication

Not applicable

### Availability of data and material

The datasets used and/or analyzed during the current study are available from the corresponding author on reasonable request.

### Authors’ contributions

Conceptualization, D.L.; methodology, D.L. and G.C.; validation, D.L., G.C. and A.S.; formal analysis, D.L., G.C.; data curation, D.L., F.S., A-E.O.; writing—original draft preparation, D.L., F.S., A.S.; writing— review and editing, D.L., G.C., S.S, A.S., A-E.O.; supervision, D.L. All authors have read and agreed to the published version of the manuscript.

## Acknowledgements

Not applicable

## Figure Legend

**Figure S1.**
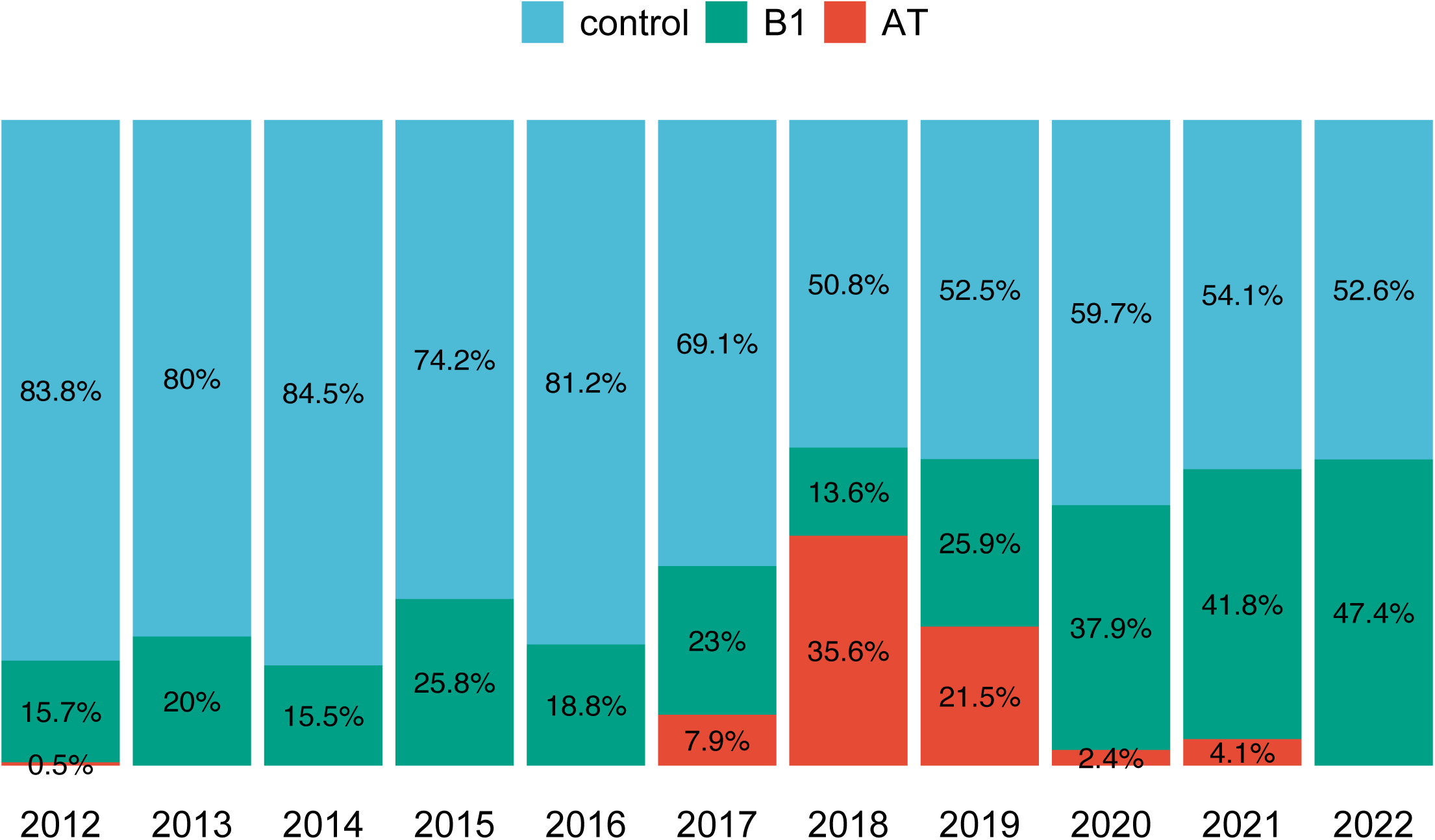
proportion of treatment group assignment along time: Stacked barplot showing the proportion of each treatment group along years of admission.

## References

1. Singer M, Deutschman CS, Seymour CW, Shankar-Hari M, Annane D, Bauer M, et al. The Third International Consensus Definitions for Sepsis and Septic Shock (Sepsis-3). JAMA. 2016;315:801.

2. Vincent J-L, Sakr Y, Sprung CL, Ranieri VM, Reinhart K, Gerlach H, et al. Sepsis in European intensive care units: results of the SOAP study. Crit Care Med. 2006;34:344–53.

3. Xie J, Wang H, Kang Y, Zhou L, Liu Z, Qin B, et al. The Epidemiology of Sepsis in Chinese ICUs: A National Cross-Sectional Survey. Crit Care Med. 2020;48:e209–18.

4. Vincent J-L, Marshall JC, Namendys-Silva SA, François B, Martin-Loeches I, Lipman J, et al. Assessment of the worldwide burden of critical illness: the intensive care over nations (ICON) audit. Lancet Respir Med. 2014;2:380–6.

5. Sakr Y, Jaschinski U, Wittebole X, Szakmany T, Lipman J, Ñamendys-Silva SA, et al. Sepsis in Intensive Care Unit Patients: Worldwide Data From the Intensive Care over Nations Audit. Open Forum Infect Dis [Internet]. 2018 [cited 2020 Nov 10];5. Available from: https://www.ncbi.nlm.nih.gov/pmc/articles/PMC6289022/

6. Fleischmann C, Scherag A, Adhikari NKJ, Hartog CS, Tsaganos T, Schlattmann P, et al. Assessment of Global Incidence and Mortality of Hospital-treated Sepsis. Current Estimates and Limitations. Am J Respir Crit Care Med. American Thoracic Society - AJRCCM; 2015;193:259–72.

7. Kaukonen K-M, Bailey M, Pilcher D, Cooper DJ, Bellomo R. Systemic Inflammatory Response Syndrome Criteria in Defining Severe Sepsis. New England Journal of Medicine. 2015;372:150319144911000.

8. Dombrovskiy VY, Martin AA, Sunderram J, Paz HL. Rapid increase in hospitalization and mortality rates for severe sepsis in the United States: a trend analysis from 1993 to 2003. Crit Care Med. 2007;35:1244–50.

9. Vincent J-L, Lefrant J-Y, Kotfis K, Nanchal R, Martin-Loeches I, Wittebole X, et al. Comparison of European ICU patients in 2012 (ICON) versus 2002 (SOAP). Intensive Care Med. 2018;44:337–44.

10. Howell MD, Davis AM. Management of Sepsis and Septic Shock. JAMA. 2017;317:847.

11. Yealy DM, Mohr NM, Shapiro NI, Venkatesh A, Jones AE, Self WH. Early Care of Adults With Suspected Sepsis in the Emergency Department and Out-of-Hospital Environment: A Consensus-Based Task Force Report. Annals of Emergency Medicine. 2021;78:1–19.

12. Evans L, Rhodes A, Alhazzani W, Antonelli M, Coopersmith CM, French C, et al. Surviving sepsis campaign: international guidelines for management of sepsis and septic shock 2021. Intensive Care Med. 2021;47:1181–247.

13. Marik PE, Khangoora V, Rivera R, Hooper MH, Catravas J. Hydrocortisone, Vitamin C, and Thiamine for the Treatment of Severe Sepsis and Septic Shock: A Retrospective Before-After Study. Chest. 2017;151:1229–38.

14. Assouline B, Faivre A, Verissimo T, Sangla F, Berchtold L, Giraud R, et al. Thiamine, Ascorbic Acid, and Hydrocortisone As a Metabolic Resuscitation Cocktail in Sepsis: A Meta-Analysis of Randomized Controlled Trials With Trial Sequential Analysis. Crit Care Med. 2021;49:2112–20.

15. Stekhoven DJ, Bühlmann P. MissForest--non-parametric missing value imputation for mixed-type data. Bioinformatics. 2012;28:112–8.

16. Hassan MM, Atiya AF, El-Gayar N, El-Fouly R. Regression in the Presence Missing Data Using Ensemble Methods. 2007 International Joint Conference on Neural Networks. 2007. p. 1261–5.

17. Nanni L, Lumini A, Brahnam S. A classifier ensemble approach for the missing feature problem. Artif Intell Med. 2012;55:37–50.

18. Jaeger BC, Cantor R, Sthanam V, Xie R, Kirklin JK, Rudraraju R. Improving Outcome Predictions for Patients Receiving Mechanical Circulatory Support by Optimizing Imputation of Missing Values. Circ Cardiovasc Qual Outcomes. 2021;14:e007071.

19. Perez-Lebel A, Varoquaux G, Le Morvan M, Josse J, Poline J-B. Benchmarking missing-values approaches for predictive models on health databases. GigaScience. 2022;11:giac013.

20. Ridgeway G, McCaffrey DF, Morral AR, Cefalu M, Burgette LF, Pane JD, et al. Toolkit for Weighting and Analysis of Nonequivalent Groups: A Tutorial for the R TWANG Package [Internet]. RAND Corporation; 2022 Jul. Available from: https://www.rand.org/pubs/tools/TLA570-5.html

21. McCaffrey DF, Griffin BA, Almirall D, Slaughter ME, Ramchand R, Burgette LF. A tutorial on propensity score estimation for multiple treatments using generalized boosted models. Statistics in Medicine. 2013;32:3388–414.

22. Stuart EA. Matching methods for causal inference: A review and a look forward. Stat Sci. 2010;25:1–21.

23. Grambsch PM, Therneau TM. Proportional hazards tests and diagnostics based on weighted residuals. Biometrika. 1994;

24. Bang H, Robins JM. Doubly robust estimation in missing data and causal inference models. Biometrics. 2005;61:962–73.

25. Nguyen T-L, Collins GS, Spence J, Daurès J-P, Devereaux PJ, Landais P, et al. Double-adjustment in propensity score matching analysis: choosing a threshold for considering residual imbalance. BMC Medical Research Methodology. 2017;17:78.

26. Rubin DB, Thomas N. Combining Propensity Score Matching with Additional Adjustments for Prognostic Covariates. Journal of the American Statistical Association. Taylor & Francis; 2000;95:573–85.

27. Fujii T, Luethi N, Young PJ, Frei DR, Eastwood GM, French CJ, et al. Effect of Vitamin C, Hydrocortisone, and Thiamine vs Hydrocortisone Alone on Time Alive and Free of Vasopressor Support Among Patients With Septic Shock: The VITAMINS Randomized Clinical Trial. JAMA. 2020;323:423–31.

28. Moskowitz A, Huang DT, Hou PC, Gong J, Doshi PB, Grossestreuer AV, et al. Effect of Ascorbic Acid, Corticosteroids, and Thiamine on Organ Injury in Septic Shock: The ACTS Randomized Clinical Trial. JAMA. 2020;324:642–50.

29. Chang P, Liao Y, Guan J, Guo Y, Zhao M, Hu J, et al. Combined Treatment With Hydrocortisone, Vitamin C, and Thiamine for Sepsis and Septic Shock: A Randomized Controlled Trial. Chest. 2020;158:174–82.

30. Wani SJ, Mufti SA, Jan RA, Shah SU, Qadri SM, Khan UH, et al. Combination of vitamin C, thiamine and hydrocortisone added to standard treatment in the management of sepsis: results from an open label randomised controlled clinical trial and a review of the literature. Infect Dis (Lond). 2020;52:271–8.

31. Iglesias J, Vassallo AV, Patel VV, Sullivan JB, Cavanaugh J, Elbaga Y. Outcomes of Metabolic Resuscitation Using Ascorbic Acid, Thiamine, and Glucocorticoids in the Early Treatment of Sepsis: The ORANGES Trial. Chest. 2020;158:164–73.

32. Mohamed ZU, Prasannan P, Moni M, Edathadathil F, Prasanna P, Menon A, et al. Vitamin C Therapy for Routine Care in Septic Shock (ViCTOR) Trial: Effect of Intravenous Vitamin C, Thiamine, and Hydrocortisone Administration on Inpatient Mortality among Patients with Septic Shock. Indian J Crit Care Med. 2020;24:653–61.

33. Reddy PR, Samavedam S, Aluru N, Yelle S, Rajyalakshmi B. Metabolic Resuscitation Using Hydrocortisone, Ascorbic Acid, and Thiamine: Do Individual Components Influence Reversal of Shock Independently? Indian J Crit Care Med. 2020;24:649–52.

34. Lyu Q-Q, Zheng R-Q, Chen Q-H, Yu J-Q, Shao J, Gu X-H. Early administration of hydrocortisone, vitamin C, and thiamine in adult patients with septic shock: a randomized controlled clinical trial. Crit Care. 2022;26:295.

35. Hussein AA, Sabry NA, Abdalla MS, Farid SF. A prospective, randomised clinical study comparing triple therapy regimen to hydrocortisone monotherapy in reducing mortality in septic shock patients. Int J Clin Pract. 2021;75:e14376.

36. Nabil T, Islam A. Early Adjuvant Intravenous Vitamin C Treatment in Septic Shock may Resolve the Vasopressor Dependence. International Journal of Microbiology & Advanced Immunology. 2017;77–81.

37. Zabet MH, Mohammadi M, Ramezani M, Khalili H. Effect of high-dose Ascorbic acid on vasopressor’s requirement in septic shock. J Res Pharm Pract. 2016;5:94–100.

38. Fowler AA, Syed AA, Knowlson S, Sculthorpe R, Farthing D, DeWilde C, et al. Phase I safety trial of intravenous ascorbic acid in patients with severe sepsis. J Transl Med. 2014;12:32.

39. Mahmoodpoor A, Shadvar K, Sanaie S, Hadipoor MR, Pourmoghaddam MA, Saghaleini SH. Effect of Vitamin C on mortality of critically ill patients with severe pneumonia in intensive care unit: a preliminary study. BMC Infect Dis. 2021;21:616.

40. Fowler AA, Truwit JD, Hite RD, Morris PE, DeWilde C, Priday A, et al. Effect of Vitamin C Infusion on Organ Failure and Biomarkers of Inflammation and Vascular Injury in Patients With Sepsis and Severe Acute Respiratory Failure: The CITRIS-ALI Randomized Clinical Trial. JAMA. 2019;322:1261– 70.

41. Rosengrave P, Spencer E, Williman J, Mehrtens J, Morgan S, Doyle T, et al. Intravenous vitamin C administration to patients with septic shock: a pilot randomised controlled trial. Crit Care. 2022;26:26.

42. Wacker DA, Burton SL, Berger JP, Hegg AJ, Heisdorffer J, Wang Q, et al. Evaluating Vitamin C in Septic Shock: A Randomized Controlled Trial of Vitamin C Monotherapy. Crit Care Med. 2022;50:e458–67.

43. Lamontagne F, Masse M-H, Menard J, Sprague S, Pinto R, Heyland DK, et al. Intravenous Vitamin C in Adults with Sepsis in the Intensive Care Unit. N Engl J Med. Massachusetts Medical Society; 2022;386:2387–98.

44. Nandhini N, Malviya D, Parashar S, Pandey C, Nath SS, Tripathi M. Comparison of the effects of vitamin C and thiamine on refractory hypotension in patients with sepsis: A randomized controlled trial. Int J Crit Illn Inj Sci. 2022;12:138–45.

45. Donnino MW, Andersen LW, Chase M, Berg KM, Tidswell M, Giberson T, et al. Randomized, Double-Blind, Placebo-Controlled Trial of Thiamine as a Metabolic Resuscitator in Septic Shock: A Pilot Study. Critical Care Medicine. 2016;44:360–7.

46. Petsakul S, Morakul S, Tangsujaritvijit V, Kunawut P, Singhatas P, Sanguanwit P. Effects of thiamine on vasopressor requirements in patients with septic shock: a prospective randomized controlled trial. BMC Anesthesiol. 2020;20:280.

47. Depeint F, Bruce WR, Shangari N, Mehta R, O’Brien PJ. Mitochondrial function and toxicity: role of B vitamins on the one-carbon transfer pathways. Chem Biol Interact. 2006;163:113–32.

48. Legouis D, Ricksten S-E, Faivre A, Verissimo T, Gariani K, Verney C, et al. Altered proximal tubular cell glucose metabolism during acute kidney injury is associated with mortality. Nat Metab. 2020;2:732–43.

49. Cruickshank AM, Telfer AB, Shenkin A. Thiamine deficiency in the critically ill. Intensive Care Med. 1988;14:384–7.

